# Exploring the Link Between Cancer Information Complexity and Understanding Medical Statistics in Online Health Information Seeking: Insights from Health Information National Trends Survey (HINTS)

**DOI:** 10.64898/2026.03.18.26348735

**Authors:** Aditya Chakraborty, Sikta Das, Amy Myat Phyo

## Abstract

**Introduction:** Understanding the factors influencing individuals’ perceptions of cancer-related information is crucial for improving public health communication. This study explores the association between perceived difficulty in understanding information related to cancer (Cancer info Hard to Understand) and concerns about the quality of cancer-related information (Concern about Cancer Info Quality) with the extent of difficulty in comprehending medical statistics information (Understanding Medical Statistics).

**Methods:** Data came from the 2022 Health Information National Trends Survey (HINTS). The cross-sectional study included 1972 participants with a response rate of 67.36% for Cancer info Hard to Understand, and 65.31% for Concern about Cancer Info Quality. We investigated the effect of Understanding Medical Statistics on Cancer info Hard to Understand, and Concern about Cancer Info Quality using univariate and multivariable logistic regression models with survey weights. The multivariable logistic regression model was adjusted for age, gender, ethnicity, marital status, education level, employment history, confidence in internet health resources, and social media. The chi-square test was used to measure the association between the predictors and the outcome.

**Results:** Individuals finding medical statistics hard to understand were more likely to be concerned regarding the quality of the cancer-related information (AOR=1.74, 95% CI: [1.20, 2.52]) and also found cancer-related information difficult to comprehend (AOR=1.89, 95% CI: [1.19, 3.00]). Also, the influence of social media on health information seeking was significantly associated with Concern about Cancer Info Quality (AOR=2.24; 95% CI: [1.33, 3.76]), and Cancer info Hard to Understand (AOR=2.84; 95% CI: [1.61, 5.03]).

**Conclusion:** This study highlights the critical role of understanding medical statistics in shaping individuals’ perceptions of cancer-related information. From an epidemiological perspective, enhancing statistical literacy is essential for making informed health decisions, addressing health disparities, and designing effective, targeted cancer communication strategies.

## Introduction

Cancer continues to be one of the foremost causes of illness and death worldwide, and its effects go beyond clinical results to include how patients perceive and understand the quality of care they receive. In a time where patient-centered healthcare is paramount, it is essential to grasp how individuals interpret cancer-related information and assess the quality of cancer treatment, as this knowledge is vital for guiding policy, enhancing health communication strategies, and minimizing cancer outcome disparities. This research explores two significant psychosocial outcomes using the Health Information National Trends Survey (HINTS): “Cancer is too difficult to understand” and “Concerns regarding the quality of cancer care.” These response variables highlight crucial aspects of cancer communication and trust within the healthcare system, especially among populations at risk of being marginalized in terms of information access.

The impetus for this research arises from a gap in literature, where most existing studies frequently focus on a singular outcome, typically associated with cancer screening habits, diagnosis, or prevention, rather than examining multiple psychosocial factors affecting cancer experiences simultaneously. Although earlier research using HINTS data has thoroughly analyzed health information-seeking behaviors and disparities in access (Finney Rutten et al., 2020; Jiang & Liu, 2021), few have concurrently evaluated how individuals’ socio-demographic, economic, and health-related attributes affect both their understanding of cancer and their worries about the quality of care. Our study aims to fill this gap by performing a joint analysis of these two outcomes through a mix of univariate and multivariate statistical methods, thus offering a more detailed view of how these constructions overlap and what factors contribute to them.

Previous research indicates that health literacy is unevenly spread across different populations, with factors such as education, income, race/ethnicity, and access to care being significant predictors of disparities in understanding health information (Bennett et al., 2009; Viswanath & Kreuter, 2007). Likewise, individuals’ evaluations of care quality are often influenced by their previous experiences with healthcare, insurance status, and communication with healthcare providers (Martinez et al., 2014). However, many of these studies tend to isolate these elements, overlooking the possibility that limited comprehension and distrust in care may coexist or exacerbate each other. By analyzing both outcomes together, our research provides a thorough examination that can aid in identifying vulnerable subgroups that may benefit from targeted interventions.

Few studies have examined how limited comprehension and concerns about the quality of the information may work together, despite recent studies showing the connections between health literacy, psychosocial distress, and structural inequities in influencing cancer outcomes (Holden et al., 2021; Abdelhadi et al., 2023). The majority of analyses still separate these categories, concentrating on either trust in healthcare systems or comprehension of health information, but seldom at once. This untidiness neglects the potential for a feedback loop between processing challenges and doubt regarding the accuracy of cancer information. This study directs these gaps by jointly examining comprehension and care-related concerns, while also incorporating newer predictors such as digital information use. This approach offers a more integrated understanding of the psychosocial dimensions of cancer experiences and situates our work within ongoing discussions of health equity and communication disparities.

A crucial and underexplored predictor of how patients interpret cancer-related information is their understanding of medical statistics (UNDMEDSTAT). This construct reflects a person’s ability to interpret probabilities, risk ratios, treatment success rates, and statistical data—elements commonly embedded in cancer communication. Prior studies have shown that low statistical numeracy is associated with misinterpretation of survival statistics and risk communication, which can, in turn, lead to greater confusion and decreased trust in care (Lipkus & Peters, 2009; Galesic & Garcia-Retamero, 2011). For instance, individuals with low numeracy are more likely to perceive cancer as a death sentence or avoid engaging with preventive care due to misunderstandings about risk. Zikmund-Fisher et al. (2008) demonstrated that people with limited statistical literacy often struggle to make informed health decisions and rely more heavily on emotion or anecdote. Moreover, systematic literature review by Garcia-Retamero and Cokely (2017) highlights that the use of transparent visual aids can mitigate the adverse effects of low numeracy in health settings. Including medical statistics literacy in our model allows for a nuanced examination of how health numeracy contributes not only to perceived difficulty in understanding cancer but also to trust in the healthcare system.

Utilizing the nationally representative HINTS dataset, we implement robust statistical techniques to investigate the relationships between demographic, socioeconomic, and medical-related predictors and the two chosen outcome variables. By taking this approach, our study contributes to a deeper understanding of how communication and perceptions of care quality shape the cancer experience. These findings are intended to support health educators, clinicians, and policymakers in developing strategies that improve patient comprehension and build trust—particularly for communities that have historically faced barriers in accessing quality care.

## Methods

### Design

In this section, we present our main methodology for estimating and inferring the association between the candidate’s ability to understand medical statistics and perceived difficulties and concerns about cancer-related information. For this paper, we obtained the data from 2022 Health Information National Trends Survey 6 (HINTS 6). The aim of HINTS is to synthesize data regarding the population’s attitudes toward health-related information, including their health behaviors, opinions, and knowledge, and is nationally representative of the American public [1]. HINTS was fielded to a random sample of noninstitutionalized adults 18 years of age and older in the United States to explore their cancer risk behaviors, health information seeking and usage, beliefs about cancer, and use of technology to manage their health [2].

### Sample

Based on the success of the mixed-method sample collection for HINTS 5, Cycle 3, HINTS 6 was offered to participants either online or on paper. HINTS 6 (2022) used a two-stage, address-based sampling design. First, a stratified random sample of residential addresses was selected from the Marketing Systems Group (MSG) national address database. Second, within each sampled household, one adult was randomly selected to participate. The sampling frame comprised all deliverable residential addresses in the MSG file. The final dataset included 6,252 completed surveys [3]. We analyzed the data using survey weighting, as it allows researchers to generalize the results to the US population. Survey weights based on population estimates from the American Community Survey account for nonresponse and coverage error. We used jackknife replicate weights to provide bias-corrected variance estimates [4]. HINTS uses 50 replicate weights, each of which is obtained by removing 1/50^th^ of the total sample and adjusting the weights accordingly (Nelson et al., 2005).

### Measures

The main two outcome variables in this study were: Cancer info Hard to Understand, which implies candidate’s perceived difficulty in understanding cancer related information and Concern about Cancer Info Quality, concerns about the quality of the cancer related information. The cross-sectional study included 1972 participants with a response rate of 67.36% for Cancer info Hard to Understand, and 65.31% for Concern about Cancer Info Quality.

The primary exposure variable of interest is Understanding Medical Statistics which captures how easy or hard participants found it to understand medical statistics (National Cancer Institute, 2022). For Understanding Medical Statistics, the responses were recoded into a binary format using SAS PROC FORMAT: responses of “Easy” and “Very easy” were merged and coded as 0 while “Hard” and “Very hard” were combined and coded as 1.

### Independent Variables

Additional demographic variables included sex at birth, recoded as a binary variable with categories male and female, age, grouped into five categories 18–34, 35–49, 50–64, 65–74, and 75 years or older, marital status as a categorical variable (Married, Living as married or with a romantic partner, Divorced, Widowed, Separated, and Single, never been married), race/ethnicity (Hispanic, Non-Hispanic Asian, Non-Hispanic Black or African American, Non-Hispanic Other, and Non-Hispanic White), household income ($20,000, $20,000 to < $35,000, $35,000 to < $50,000, $50,000 to < $75,000, and $75,000 or more), employment status was assessed with a binary variable indicating whether the respondent was employed full-time (Yes or No), history of cancer captured personal history of cancer diagnosis, was included as a binary variable (Yes/No), health insurance as a binary indicator of current insurance coverage (Yes/No). Additionally, two psychological variables confidence in online health and difficult to interpret social media info were included captures candidates’ feelings by asking them questions like “How confident are you that you can find helpful health resources on the Internet” and responses range from “Completely confident”, “Very confident”, “Somewhat confident”, “A little confident”, “Not confident at all”. Candidate’s agreeing on the level of difficulty in acquiring the health information from social media is obtained by the variable difficult to interpret social media info with its responses “Strongly disagree”, “Somewhat disagree”, “Somewhat agree”, and “Strongly agree.

### Statistical Analysis

The data cleaning part was performed in (RStudio Team, 2022). The dataset initially consisted of 6,252 observations. To ensure accuracy and integrity of the analysis, observations with missing or invalid data - such as responses labeled “Missing data (Not Ascertained)”, “Missing data (Web partial - Question Never Seen)”, “Multiple responses selected in error”, “Unreadable or Nonconforming Numeric Response” - on any study variable were excluded. After this step, a final analytical sample of 1,972 participants was retained for both univariate and multivariate analyses, which were performed on SAS software 9.4 (SAS Institute Inc., 2021).

We performed univariate and multivariate analysis using PROC SURVEYLOGISTIC. We ran separate logistic regression model for each categorical predictor including age group, sex at birth, marital status, race/ethnicity, education, household income, employment status, health insurance, history of cancer diagnosis, confidence in internet health information (Confidence in online health), use of social media (Difficult to Interpret Social Media Info), and perceived difficulty understanding medical statistics (Understanding Medical Statistics). For the regression analysis, categorical variables were transformed into binary variables using SAS proc format. Chi-square test was used to access the association between the predictors and the outcomes. For the outcome variables, Cancer info Hard to Understand, and Concern about Cancer Info Quality, “Strongly agree” and “Somewhat agree” were coded as 1 (indicating agreement), while “Somewhat disagree” and “Strongly disagree” were recoded as 0 (indicating disagreement).

Univariate, and multivariable logistic regression models were then fitted for both the outcome variables, adjusting for all the covariates listed above. For the categorical variables, we specified the references to allow for meaningful comparisons and applied appropriate formats to dichotomize responses for Understanding Medical Statistics, Concern about Cancer Info Quality and Cancer info Hard to Understand.

The models generated adjusted odds ratios (AORs) and 95% confidence intervals (CI) to quantify associations between the predictors and outcomes. All analyses were conducted using a statistical significance level (α) at 0.05.

## Results

Data from 6,252 survey respondents were used. After eliminating observations with missing, invalid data, 1,972 participants were retained for further analyses. In Table 1 and Table 2, we present the weighted frequency tables and association between the covariates and the outcome variables Cancertoohardunderstand and Cancerconcernedquality respectively, their percentage estimates, and the corresponding 95% confidence intervals, featuring the demographic characteristics.

**Table 1:** Bivariate Associations Between Participant Characteristics and Cancer Info Hard to Understand.

| Variables | Cancer info Hard to Understand |  |  |  |  |  |
| --- | --- | --- | --- | --- | --- | --- |
|  | Overall (N=1972) |  | Agree (N=706) |  | Disagree (N=1266) |  |
|  | N | Weighted %<br>(95% CI) | N | Weighted %<br>(95% CI) | N | Weighted %<br>(95% CI) |
| **Primary Exposure** |  |  |  |  |  |  |
| **Understanding Medical Statistics** |  |  |  |  |  |  |
| Easy | 1554 | 78.74 (75.42-82.06) | 460 | 25.40 (22.60-28.21) | 1094 | 53.34 (49.99-56.69) |
| Hard | 418 | 21.26 (17.93-24.58) | 246 | 11.57 (9.01-14.14) | 172 | 9.68 (7.33-12.02) |
| **Covariates** |  |  |  |  |  |  |
| **Sex at Birth** |  |  |  |  |  |  |
| Male | 691 | 38.30 (35.55-41.04) | 252 | 14.88 (12.06-17.72) | 439 | 23.41 (21.07-25.75) |
| Female | 1281 | 61.70 (58.95-64.44) | 454 | 22.09 (19.25-24.93) | 827 | 39.61 (36.75-42.46) |
| **Marital Status** |  |  |  |  |  |  |
| Divorced | 284 | 6.45 (5.17-7.72) | 99 | 2.25 (1.62-2.89) | 185 | 4.19 (3.26-5.12) |
| Living as married or with partner | 154 | 6.19 (4.79-7.58) | 59 | 2.07 (1.42-2.71) | 95 | 4.12 (2.97-5.27) |
| Married | 1017 | 58.59 (55.63-61.55) | 348 | 21.08 (18.24-23.92) | 669 | 37.51 (34.64-40.38) |
| Separated | 30 | 0.94 (0.40-1.47) | 10 | 0.22 (0.07-0.36) | 20 | 0.72 (0.23-1.21) |
| Single, never married | 348 | 24.89 (21.77-28.01) | 117 | 9.55 (6.79-12.31) | 231 | 15.34 (12.65-18.03) |
| Widowed | 139 | 2.95 (2.11-3.79) | 73 | 1.81 (1.17-2.46) | 66 | 1.14 (0.72-1.55) |
| **Age-group** |  |  |  |  |  |  |
| 18-34 | 341 | 24.35 (20.84-27.86) | 139 | 10.39 (6.98-13.79) | 202 | 13.96 (11.47-16.45) |
| 35-49 | 469 | 28.70 (25.36-32.05) | 170 | 9.36 (7.26-11.45) | 299 | 19.35 (16.57-22.12) |
| 50-64 | 598 | 30.21 (27.12- | 198 | 10.67 (7.98- | 400 | 19.54 (16.55-22.53) |
|  |  | 33.31) |  | 13.37) |  |  |
| 65-74 | 398 | 11.77 (10.13-13.42) | 136 | 4.70 (3.53-5.86) | 262 | 7.08 (5.89-8.26) |
| 75+ | 166 | 4.96 (4.01-5.91) | 63 | 1.87 (1.25-2.49) | 103 | 3.09 (2.40-3.79) |
| **Race/ethnicity** |  |  |  |  |  |  |
| Hispanic | 281 | 13.64 (11.47-15.80) | 112 | 5.77 (4.03-7.51) | 169 | 7.86 (5.83-9.90) |
| Non-Hispanic Asian | 103 | 5.11 (3.71-6.51) | 44 | 2.37 (1.24-3.49) | 59 | 2.74 (1.68-3.81) |
| Non-Hispanic Black or African American | 219 | 8.39 (6.64-10.15) | 66 | 2.18 (1.06-3.30) | 153 | 6.21 (4.87-7.56) |
| Non-Hispanic Other | 74 | 4.98 (3.06-6.90) | 38 | 3.08 (1.28-4.87) | 36 | 1.91 (0.92-2.89) |
| Non-Hispanic White | 1295 | 67.88 (65.20-70.56) | 446 | 23.58 (20.44-26.72) | 849 | 44.30 (40.89-47.70) |
| **Household Income** |  |  |  |  |  |  |
| Less than \$20,000 | 175 | 8.57 (5.84-11.30) | 73 | 3.91 (1.96-5.86) | 102 | 4.66 (3.07-6.25) |
| \$20,000 to < \$35,000 | 189 | 6.64 (5.26-8.03) | 63 | 2.22 (1.48-2.96) | 126 | 4.43 (3.26-5.12) |
| \$35,000 to < \$50,000 | 216 | 10.04 (7.87-12.22) | 95 | 4.17 (2.93-5.40) | 121 | 5.88 (4.34-7.41) |
| \$50,000 to < \$75,000 | 377 | 17.78 (15.38-20.18) | 134 | 6.91 (5.21-8.62) | 243 | 10.87 (8.73-13.00) |
| \$75,000 or More | 1015 | 56.96 (53.27-60.65) | 341 | 19.78 (16.75-22.80) | 674 | 37.19 (34.42-39.95) |
| **Education** |  |  |  |  |  |  |
| Less than High School | 56 | 2.81 (1.68-3.93) | 23 | 0.80 (0.41-1.19) | 33 | 2.01 (0.84-3.17) |
| High School Graduate | 198 | 12.70 (10.17-15.22) | 80 | 5.32 (3.10-7.55) | 118 | 7.37 (5.55-9.20) |
| Some College | 524 | 39.36 (36.08-42.64) | 207 | 15.95 (12.98-18.92) | 317 | 23.41 (20.10-26.72) |
| College Graduate or More | 1194 | 45.14 (42.55-47.73) | 396 | 14.91 (12.94-16.88) | 798 | 30.23 (27.84-32.62) |
| **Employment status** |  |  |  |  |  |  |
| No | 897 | 42.27 (38.37-46.17) | 314 | 15.65 (12.88-18.42) | 583 | 26.62 (23.19-30.05) |
| Yes | 1075 | 57.73 (53.83-61.63) | 392 | 21.33 (18.74-23.92) | 683 | 36.40 (32.56-40.23) |
| **History of cancer** |  |  |  |  |  |  |
| No | 1584 | 85.45 (83.87-87.03) | 585 | 32.37 (28.97-35.77) | 999 | 53.08 (49.73-56.42) |
| Yes | 388 | 14.55 (12.97-16.13) | 121 | 4.61 (3.46-5.75) | 267 | 9.94 (8.40-11.48) |
| **Health Insurance** |  |  |  |  |  |  |
| No | 121 | 8.27 (5.92-10.63) | 49 | 3.00 (1.49-4.51) | 72 | 5.27 (3.13-7.41) |
| Yes | 1851 | 91.73 (89.37- | 657 | 33.98 (30.88- | 1194 | 57.75 (54.41-61.09) |
|  |  | 94.08) |  | 37.07) |  |  |
| **Confidence in online health** |  |  |  |  |  |  |
| Not confident at all | 25 | 1.08 (0.52-1.64) | 13 | 0.56 (0.19-0.93) | 12 | 0.52 (0.06-0.99) |
| A little confident | 119 | 5.54 (4.26-6.83) | 71 | 3.59 (2.36-4.82) | 48 | 1.95 (1.19-2.71) |
| Somewhat confident | 767 | 38.96 (35.55-42.38) | 351 | 18.66 (15.79-21.54) | 416 | 20.30 (17.24-23.36) |
| Very confident | 783 | 39.07 (35.19-42.95) | 226 | 11.44 (9.14-13.74) | 557 | 27.63 (24.46-30.81) |
| Completely confident | 278 | 15.34 (12.74-17.95) | 45 | 2.73 (1.69-3.76) | 233 | 12.61 (10.08-15.14) |
| **Difficult to Interpret Social Media Info** |  |  |  |  |  |  |
| Strongly disagree | 297 | 14.22 (11.94-16.49) | 76 | 2.94 (2.09-3.79) | 221 | 11.28 (9.01-13.54) |
| Somewhat disagree | 383 | 19.27 (16.21-22.34) | 112 | 5.60 (3.70-7.51) | 271 | 13.67 (11.34-16.00) |
| Strongly agree | 554 | 30.18 (27.10-33.26) | 259 | 15.47 (12.29-18.66) | 295 | 14.71 (12.11-17.31) |
| Somewhat agree | 738 | 36.32 (33.10-39.55) | 259 | 12.96 (11.05-14.87) | 479 | 23.36 (20.45-26.28) |

**Table 2:** Bivariate Associations Between Participant Characteristics and Concern about Cancer Info Quality.

| Variables | Concern about Cancer Info Quality |
| --- | --- |

|  | Overall (N=1972) |  | Agree (N=1098) |  | Disagree (N=874) |  |
| --- | --- | --- | --- | --- | --- | --- |
|  | N | Weighted %<br>(95% CI) | N | Weighted %<br>(95% CI) | N | Weighted %<br>(95% CI) |
| **Primary Exposure** |  |  |  |  |  |  |
| **Understanding Medical Statistics** |  |  |  |  |  |  |
| Easy | 155<br>4 | 78.74 (75.42-82.07) | 819 | 41.78 (38.28-45.28) | 735 | 36.97 (33.25-40.68) |
| Hard | 418 | 21.26 (17.93-24.58) | 279 | 14.91 (11.98-17.83) | 139 | 6.35 (4.70-8.00) |
| **Covariates** |  |  |  |  |  |  |
| **Sex at Birth** |  |  |  |  |  |  |
| Male | 691 | 38.30 (35.55-41.05) | 393 | 23.18 (19.56-26.81) | 298 | 15.12 (12.68-17.56) |
| Female | 128<br>1 | 61.70 (58.95-64.45) | 705 | 33.50 (30.23-36.77) | 576 | 28.20 (25.20-31.20) |
| **Marital Status** |  |  |  |  |  |  |
| Divorced | 284 | 6.45 (5.17-7.72) | 147 | 3.48 (2.67-4.30) | 137 | 2.96 (2.08-3.85) |
| Living as married or with partner | 154 | 6.19 (4.79-7.58) | 99 | 3.61 (2.48-4.75) | 55 | 2.57 (1.68-3.46) |
| Married | 101<br>7 | 58.59 (55.63-61.55) | 564 | 33.73 (30.09-37.36) | 453 | 24.86 (21.65-28.08) |
| Separated | 30 | 0.94 (0.40-1.47) | 13 | 0.26 (0.11-0.41) | 17 | 0.68 (0.18-1.17) |
| Single, never married | 348 | 24.89 (21.77-28.01) | 189 | 13.66 (10.57-16.75) | 159 | 11.23 (8.78-13.69) |
| Widowed | 139 | 2.95 (2.11-3.79) | 86 | 1.94 (1.38-2.51) | 53 | 1.01 (0.46-1.56) |
| **Age-group** |  |  |  |  |  |  |
| 18-34 | 341 | 24.35 (20.84-27.86) | 208 | 14.14 (10.45-17.83) | 133 | 10.21 (7.85-12.57) |
| 35-49 | 469 | 28.70 (25.36-32.05) | 273 | 15.73 (13.15-18.31) | 196 | 12.97 (10.28-15.67) |
| 50-64 | 598 | 30.21 (27.12-33.31) | 332 | 18.43 (15.32-21.53) | 266 | 11.79 (9.41-14.17) |
| 65-74 | 398 | 11.77 (10.13-13.42) | 202 | 5.96 (4.80-7.12) | 196 | 5.81 (4.67-6.95) |
| 75+ | 166 | 4.96 (4.01-5.91) | 83 | 2.43 (1.65-3.20) | 83 | 2.54 (1.91-3.17) |
| **Race/ethnicity** |  |  |  |  |  |  |
| Hispanic | 281 | 13.64 (11.47-15.80) | 157 | 8.07 (6.28-9.87) | 124 | 5.56 (3.89-7.24) |
| Non-Hispanic Asian | 103 | 5.11 (3.71-6.51) | 64 | 3.33 (2.05-4.61) | 39 | 1.78 (0.96-2.59) |
| Non-Hispanic Black or African American | 219 | 8.39 (6.64-10.15) | 103 | 3.71 (2.38-5.04) | 116 | 4.69 (3.31-6.07) |
| Non-Hispanic Other | 74 | 4.98 (3.06-6.90) | 50 | 3.85 (2.00-5.70) | 24 | 1.13 (0.34-1.93) |
| Non-Hispanic White | 129 | 67.88 (65.20- | 724 | 37.72 (34.42- | 571 | 30.16 (26.57- |
|  | 5 | 70.56) |  | 41.02) |  | 33.75) |
| **Household Income** |  |  |  |  |  |  |
| Less than \$20,000 | 175 | 8.57 (5.84-11.30) | 94 | 4.50 (2.60-6.39) | 81 | 4.07 (2.34-5.81) |
| \$20,000 to < \$35,000 | 189 | 6.64 (5.26-8.03) | 109 | 4.02 (2.98-5.06) | 80 | 2.63 (1.71-3.54) |
| \$35,000 to < \$50,000 | 216 | 10.04 (7.87-12.22) | 115 | 4.83 (3.23-6.43) | 101 | 5.21 (3.66-6.77) |
| \$50,000 to < \$75,000 | 377 | 17.78 (15.38-20.18) | 199 | 9.66 (7.96-11.36) | 178 | 8.12 (6.24-9.99) |
| \$75,000 or More | 101<br>5 | 56.96 (53.27-60.65) | 581 | 33.67 (29.50-37.85) | 434 | 23.29 (19.83-26.74) |
| **Education** |  |  |  |  |  |  |
| Less than High School | 56 | 2.81 (1.68-3.93) | 29 | 1.28 (0.54-2.02) | 27 | 1.53 (0.56-2.50) |
| High School Graduate | 198 | 12.70 (10.17-15.22) | 98 | 6.94 (4.40-9.48) | 100 | 5.76 (4.08-7.43) |
| Some College | 524 | 39.36 (36.08-42.64) | 298 | 22.21 (18.54-25.89) | 226 | 17.14 (14.16-20.12) |
| College Graduate or More | 119<br>4 | 45.14 (42.55-47.73) | 673 | 26.25 (23.67-28.83) | 521 | 18.89 (16.38-21.41) |
| **Employment status** |  |  |  |  |  |  |
| No | 897 | 42.27 (38.37-46.17) | 483 | 22.83 (19.90-25.77) | 414 | 19.43 (16.81-22.06) |
| Yes | 107<br>5 | 57.73 (53.83-61.63) | 615 | 33.85 (30.26-37.44) | 460 | 23.88 (20.30-27.46) |
| **History of cancer** |  |  |  |  |  |  |
| No | 158<br>4 | 85.45 (83.87-87.03) | 901 | 48.92 (44.89-52.96) | 683 | 36.53 (32.95-40.10) |
| Yes | 388 | 14.55 (12.97-16.13) | 197 | 7.76 (6.58-8.93) | 191 | 6.79 (5.55-8.03) |
| **Health Insurance** |  |  |  |  |  |  |
| No | 121 | 8.27 (5.92-10.63) | 72 | 4.84 (3.04-6.64) | 49 | 3.43 (1.73-5.13) |
| Yes | 185<br>1 | 91.73 (89.37-94.08) | 102<br>6 | 51.84 (48.05-55.63) | 825 | 39.89 (36.20-43.57) |
| **Confidence in online health** |  |  |  |  |  |  |
| Not confident at all | 25 | 1.08 (0.52-1.64) | 17 | 0.79 (0.29-1.29) | 8 | 0.29 (0.00-0.58) |
| A little confident | 119 | 5.54 (4.26-6.83) | 89 | 3.86 (2.80-4.91) | 30 | 1.69 (0.80-2.58) |
| Somewhat confident | 767 | 38.96 (35.55-42.38) | 509 | 26.74 (23.63-29.85) | 258 | 12.22 (9.87-14.58) |
| Very confident | 783 | 39.07 (35.19-42.95) | 390 | 20.15 (16.78-23.52) | 393 | 18.93 (16.36-21.50) |
| Completely confident | 278 | 15.34 (12.74-17.95) | 93 | 5.15 (3.77-6.54) | 185 | 10.19 (7.46-12.92) |

| **Difficult to Interpret Social Media Info** |  |  |  |  |  |  |
| --- | --- | --- | --- | --- | --- | --- |
| Strongly disagree | 297 | 14.22 (11.94-16.49) | 123 | 6.23 (4.54-7.93) | 174 | 7.98 (5.97-10.00) |
| Somewhat disagree | 383 | 19.27 (16.21-22.34) | 200 | 9.99 (7.52-12.47) | 183 | 9.28 (7.07-11.49) |
| Strongly agree | 554 | 30.18 (27.10-33.26) | 361 | 20.05 (16.81-23.30) | 193 | 10.13 (8.14-12.12) |
| Somewhat agree | 738 | 36.32 (33.10-39.55) | 414 | 20.40 (17.71-23.10) | 324 | 15.92 (13.29-18.55) |

The sample consists of a weighted majority (61.70%) of female respondents and 38.30% of male respondents. Majority were found to be married (weighted percentage: 58.59%), followed by those who were single and had never married (24.89%). Smaller proportions were divorced (6.45%), cohabiting with a partner (6.19%), widowed (2.95%), and separated (0.94%).

Looking at the age distribution, the sample is skewed towards the age group 50-64 with a weighted percentage of 30.21, followed by 35-49 (28.70%), 18-34 (24.35%). Older adults aged 65–74 years comprised 11.77% of the sample, while 4.96% comprised individuals aged 75 years and above.

For the other socioeconomic variables, the majority of the sample is represented by non-Hispanic white (67.88%), followed by Hispanic (13.64%), Non-Hispanic Black or African American (8.39%), Non-Hispanic Asian (5.11%), and Non-Hispanic Other (4.98%).

In terms of annual household income, more than half (56.9%) of the weighted sample reported a combined annual income of $75,000 or above. On the other hand, 25% reported a combined annual income of less than $50,000.

For the educational background of the respondents, most of them have attended college or higher education, with a relatively very low fraction of the sample (2.81%) reporting Less than High School. In terms of employment, 57.73% are said to be employed (worked for 35 hours or more per week, for the last 30 days). The majority of the respondents (85.45%) never had cancer. It is interesting to note that most participants worried about cancer information quality regardless of their cancer diagnosis status. The majority of participants without cancer history (85.45% of the sample) expressed concerned about the quality of perceived information at 48.92%. The proportion of cancer survivors (14.55% of the sample) who expressed such concerns was 7.76%. Our data suggests varying level of confidence in finding helpful health resources on the internet such as: 39% are ‘Very confident’, 15.34% are ‘completely confident’ while 1.08% are ‘Not confident at all’. Regarding the perceived reliability on health-related information on social media, 36.3% report ‘Somewhat disagree’ that it is hard to tell if the information is true or false.

Table 1 summarizes the association between Cancertoohardunderstand and covariates, with response distributions for agreement and disagreement. 78.74% of the weighted sample reported that it is easy to interpret medical statistics. Among this group, 25.40% agree that it is hard to understand information related to cancer, while 53.34% disagree. On the other hand, 21.26% of the weighted sample finds it hard to understand medical statistics. Among this group, 11.57% agree that cancer-related information is hard to understand while 9.68% disagree. Looking at the other covariates, female respondents disagree more (39.6%) than male respondents (23.4%). Among those who agreed, 22.09% are females and 14.88% are males.

Looking at the marital status, among those who are married, 21.1% agree and 37.5% disagree to the statement, followed by other respondents single, never married. Age patterns showed that agreement peaked among 50–64-year-olds (10.67%), while disagreement was highest among 35–64-year-olds (≈19%) and lowest in adults aged 75+.

In terms of race, non-Hispanic Whites accounted for the majority of both agreement (23.58%) and disagreement (44.30%), compared to other groups. Higher income (≥$75,000) and higher education (college or more) groups also comprised the largest proportions of both “agree” and “disagree” that cancer related information is hard to understand.

In terms of education and work, college graduates (14.91% agree; 30.23% disagree), full time workers (21.33% agree; 36.40% disagree) account for a larger share of responses compared to other groups.

By health status, among those with a prior cancer diagnosis, 4.6% agree and 9.9% disagreed that cancer related information is hard to understand. Those who did not have cancer were more likely to endorse both the responses (33.98% agree; 57.75% disagree).

Most respondents were insured (91.7%), and account for the largest proportions of both agreement (33.98%) and disagreement (57.75%) that cancer-related information is too hard to understand. Confidence in internet health information showed a clear gradient: those who are very confident in finding health information on internet are more likely to disagree (27.6%), while agreement was highest among the somewhat confident group (18.6%). Perceptions of social media also shaped responses; agreement was concentrated among those who strongly (15.5%) or somewhat agreed (13.0%) that they find it hard to tell if health information on social media is true or false.

Similarly, Table 2 presents the association between the outcome variable Cancerconcernedquality and the covariates and the distribution of responses of “agree” and “disagree” responses to the statement that based on the recent research for cancer-related information, whether the respondent was concerned about the quality of the information. 78.74% of the weighted sample reported that it is easy to interpret medical statistics. Within this group, 41.78% agreed and 36.97% disagreed that they were concerned about the quality of cancer information. In contrast, among the 21.26% who found statistics hard to interpret, 14.91% agreed and 6.35% disagreed.

In terms of gender, females contributed more to both agreement (33.5%) and disagreement (28.2%) compared to males (23.2% and 15.1%). Agreement was most common among married (33.7%) and 50–64-year-old respondents (18.4%), while disagreement was highest among 35–64-year-olds (∼12–13%) and lowest among adults 75+. Non-Hispanic Whites comprised the majority of responses (37.7% agree; 30.2% disagree), while other groups accounted for smaller shares.

Socioeconomic patterns showed that those with higher income (≥$75,000) and higher education (college or more), full time workers represented the greater proportions of both agreement and disagreement compared to their complementary groups. By health status, respondents without prior cancer diagnosis, were more likely to agree (48.9%) or disagree (36.5%) than those with cancer (7.8% and 6.8%). Most respondents were insured (91.7%) and accounted for the bulk of both agreement and disagreement.

Confidence in internet health information revealed a gradient: somewhat confident (26.7%) and very confident (20.2%) respondents contributed the largest shares of agreement, while disagreement was more frequent among the very (18.9%) and completely confident (10.2%). Perceptions of social media also shaped responses, with agreement concentrated among those who strongly (20.1%) or somewhat agreed (20.4%) that it is difficult to judge health information on social media, and disagreement highest among the somewhat agreeing group (15.9%).

Table 3 and 4 illustrates the results of the univariate and multivariate analyses for the outcome variables Cancertoohardunderstand and Cancerconcernedquality.

**Table 3:**
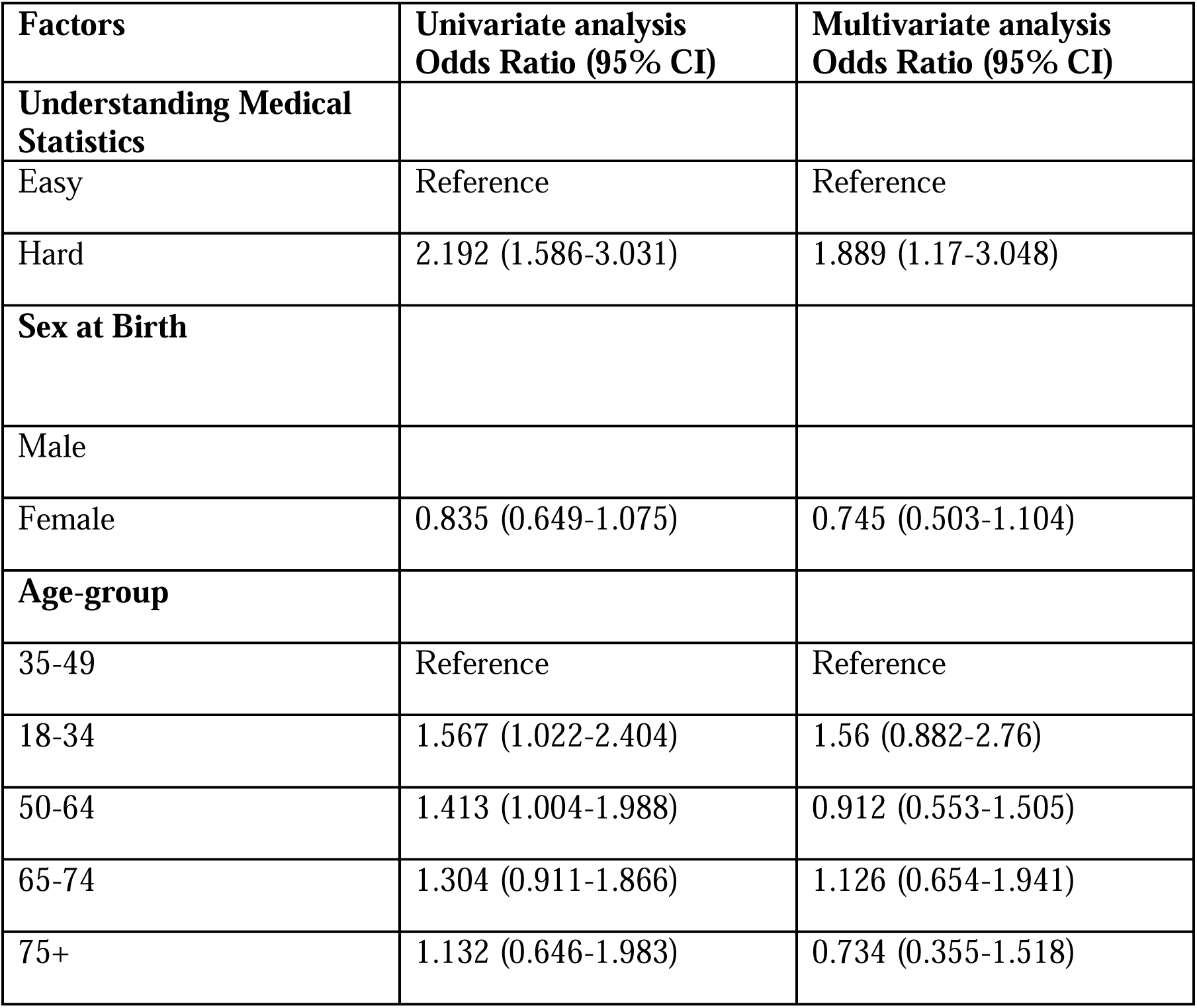
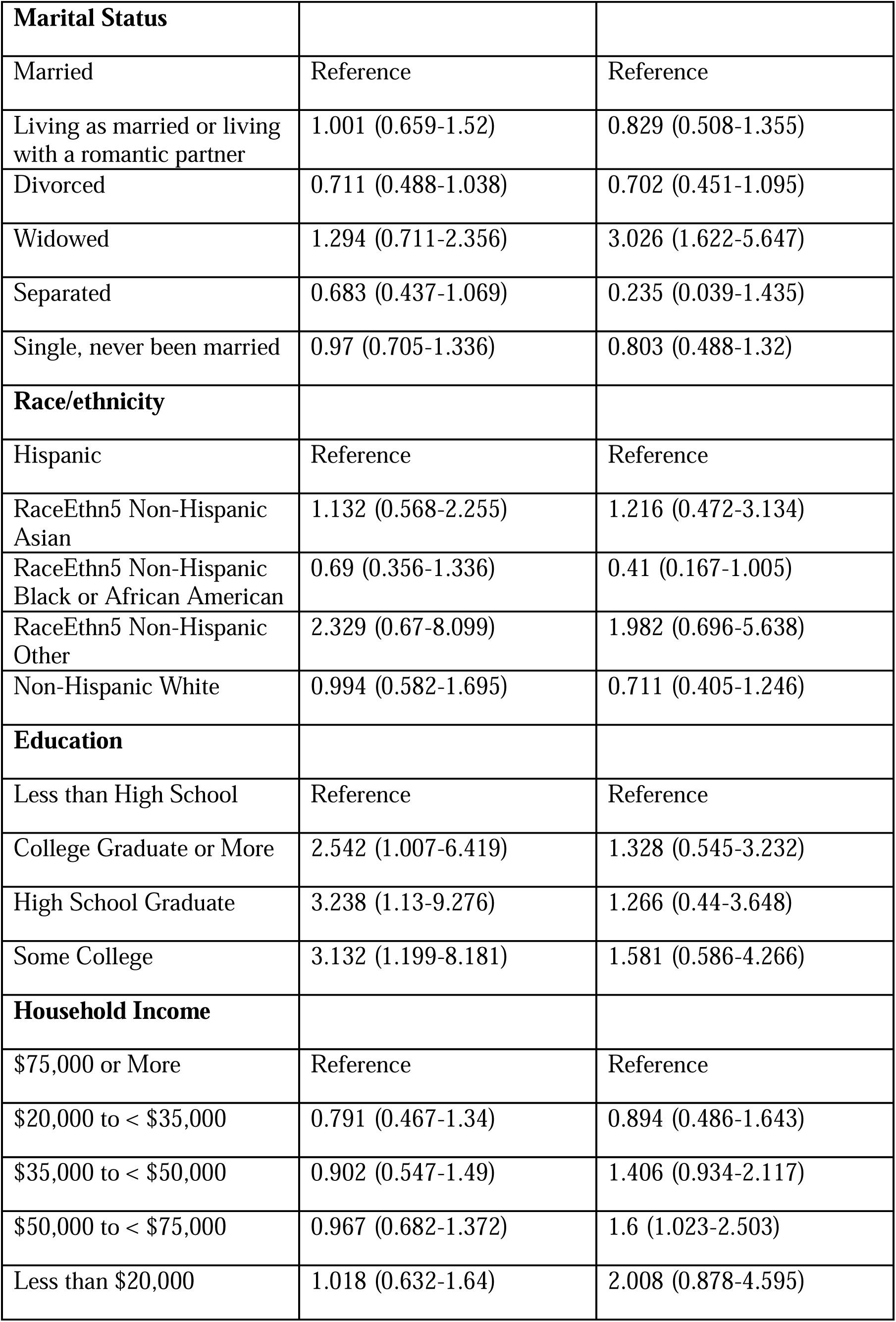
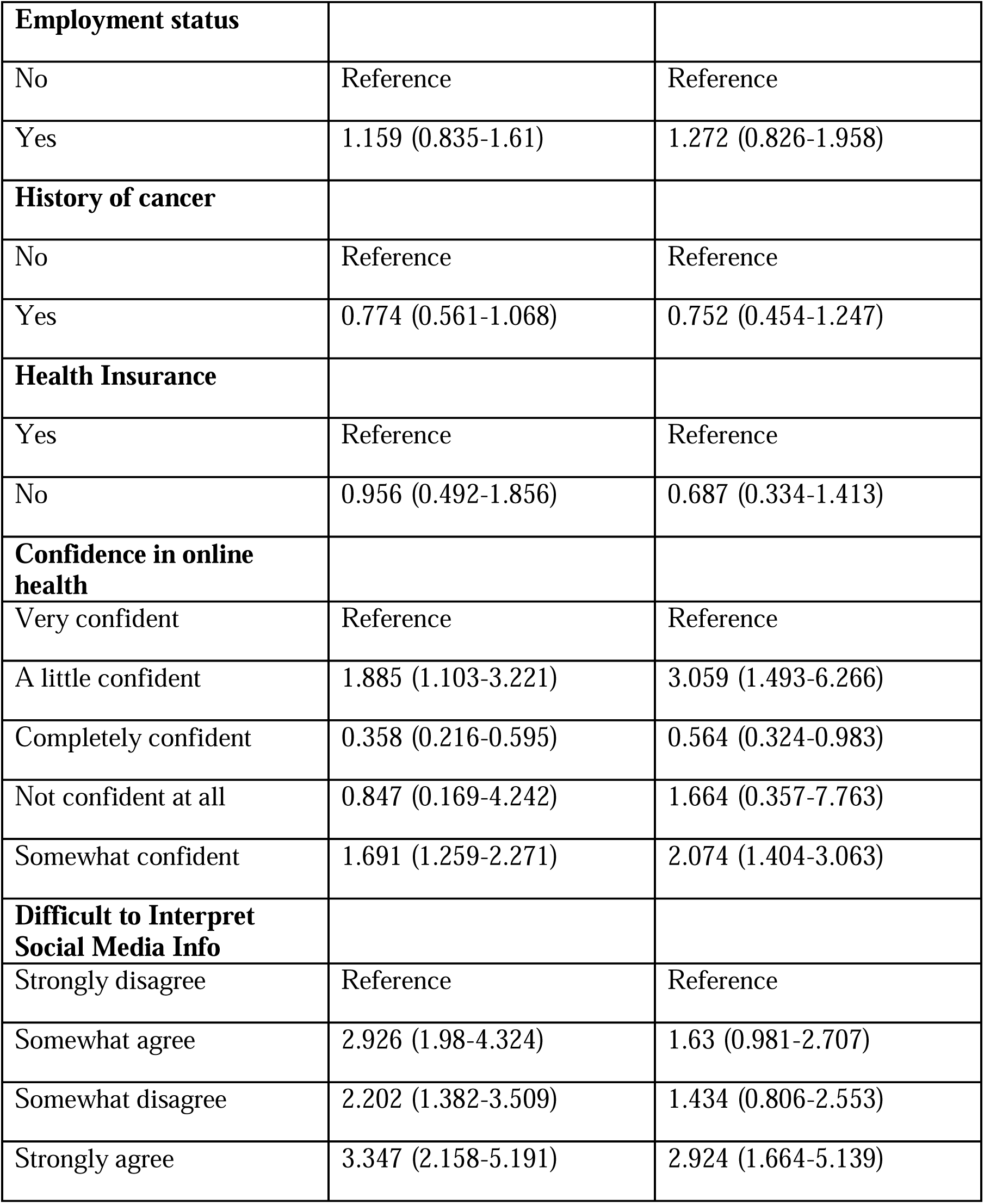
Univariate and Adjusted Multivariable Regression Analysis of the Association between Cancer Info Hard to Understand, and Participant Characteristics.

| **Factors** | **Univariate analysis**<br>**Odds Ratio (95% CI)** | **Multivariate analysis**<br>**Odds Ratio (95% CI)** |
| --- | --- | --- |
| **Understanding Medical Statistics** |  |  |
| Easy | Reference | Reference |
| Hard | 2.192 (1.586-3.031) | 1.889 (1.17-3.048) |
| **Sex at Birth** |  |  |
| Male |  |  |
| Female | 0.835 (0.649-1.075) | 0.745 (0.503-1.104) |
| **Age-group** |  |  |
| 35-49 | Reference | Reference |
| 18-34 | 1.567 (1.022-2.404) | 1.56 (0.882-2.76) |
| 50-64 | 1.413 (1.004-1.988) | 0.912 (0.553-1.505) |
| 65-74 | 1.304 (0.911-1.866) | 1.126 (0.654-1.941) |
| 75+ | 1.132 (0.646-1.983) | 0.734 (0.355-1.518) |
| **Marital Status** |  |  |
| Married | Reference | Reference |
| Living as married or living with a romantic partner | 1.001 (0.659-1.52) | 0.829 (0.508-1.355) |
| Divorced | 0.711 (0.488-1.038) | 0.702 (0.451-1.095) |
| Widowed | 1.294 (0.711-2.356) | 3.026 (1.622-5.647) |
| Separated | 0.683 (0.437-1.069) | 0.235 (0.039-1.435) |
| Single, never been married | 0.97 (0.705-1.336) | 0.803 (0.488-1.32) |
| **Race/ethnicity** |  |  |
| Hispanic | Reference | Reference |
| RaceEthn5 Non-Hispanic Asian | 1.132 (0.568-2.255) | 1.216 (0.472-3.134) |
| RaceEthn5 Non-Hispanic Black or African American | 0.69 (0.356-1.336) | 0.41 (0.167-1.005) |
| RaceEthn5 Non-Hispanic Other | 2.329 (0.67-8.099) | 1.982 (0.696-5.638) |
| Non-Hispanic White | 0.994 (0.582-1.695) | 0.711 (0.405-1.246) |
| **Education** |  |  |
| Less than High School | Reference | Reference |
| College Graduate or More | 2.542 (1.007-6.419) | 1.328 (0.545-3.232) |
| High School Graduate | 3.238 (1.13-9.276) | 1.266 (0.44-3.648) |
| Some College | 3.132 (1.199-8.181) | 1.581 (0.586-4.266) |
| **Household Income** |  |  |
| \$75,000 or More | Reference | Reference |
| \$20,000 to < \$35,000 | 0.791 (0.467-1.34) | 0.894 (0.486-1.643) |
| \$35,000 to < \$50,000 | 0.902 (0.547-1.49) | 1.406 (0.934-2.117) |
| \$50,000 to < \$75,000 | 0.967 (0.682-1.372) | 1.6 (1.023-2.503) |
| Less than \$20,000 | 1.018 (0.632-1.64) | 2.008 (0.878-4.595) |
| **Employment status** |  |  |
| No | Reference | Reference |
| Yes | 1.159 (0.835-1.61) | 1.272 (0.826-1.958) |
| **History of cancer** |  |  |
| No | Reference | Reference |
| Yes | 0.774 (0.561-1.068) | 0.752 (0.454-1.247) |
| **Health Insurance** |  |  |
| Yes | Reference | Reference |
| No | 0.956 (0.492-1.856) | 0.687 (0.334-1.413) |
| **Confidence in online health** |  |  |
| Very confident | Reference | Reference |
| A little confident | 1.885 (1.103-3.221) | 3.059 (1.493-6.266) |
| Completely confident | 0.358 (0.216-0.595) | 0.564 (0.324-0.983) |
| Not confident at all | 0.847 (0.169-4.242) | 1.664 (0.357-7.763) |
| Somewhat confident | 1.691 (1.259-2.271) | 2.074 (1.404-3.063) |
| **Difficult to Interpret Social Media Info** |  |  |
| Strongly disagree | Reference | Reference |
| Somewhat agree | 2.926 (1.98-4.324) | 1.63 (0.981-2.707) |
| Somewhat disagree | 2.202 (1.382-3.509) | 1.434 (0.806-2.553) |
| Strongly agree | 3.347 (2.158-5.191) | 2.924 (1.664-5.139) |

**Table 4:**
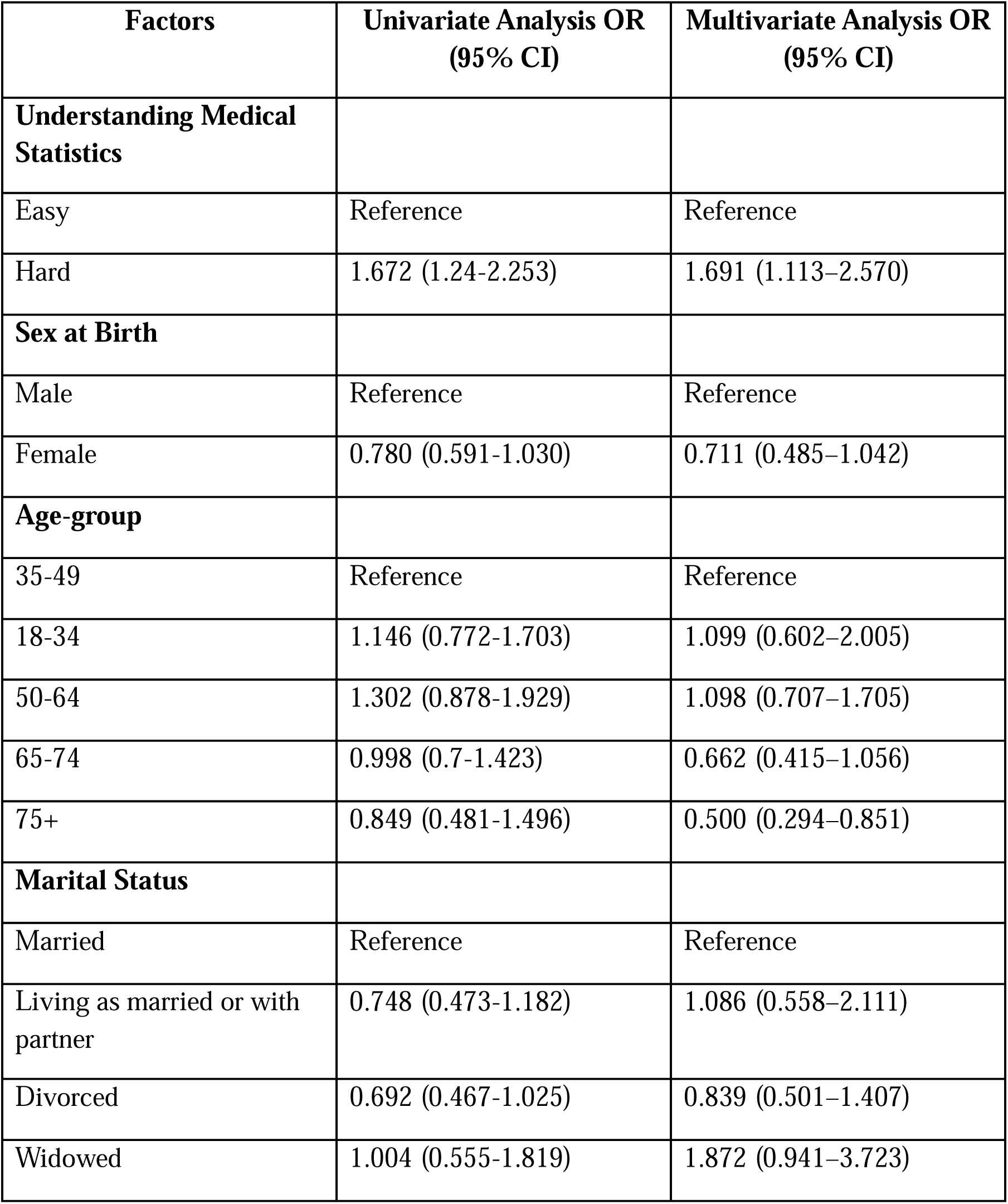
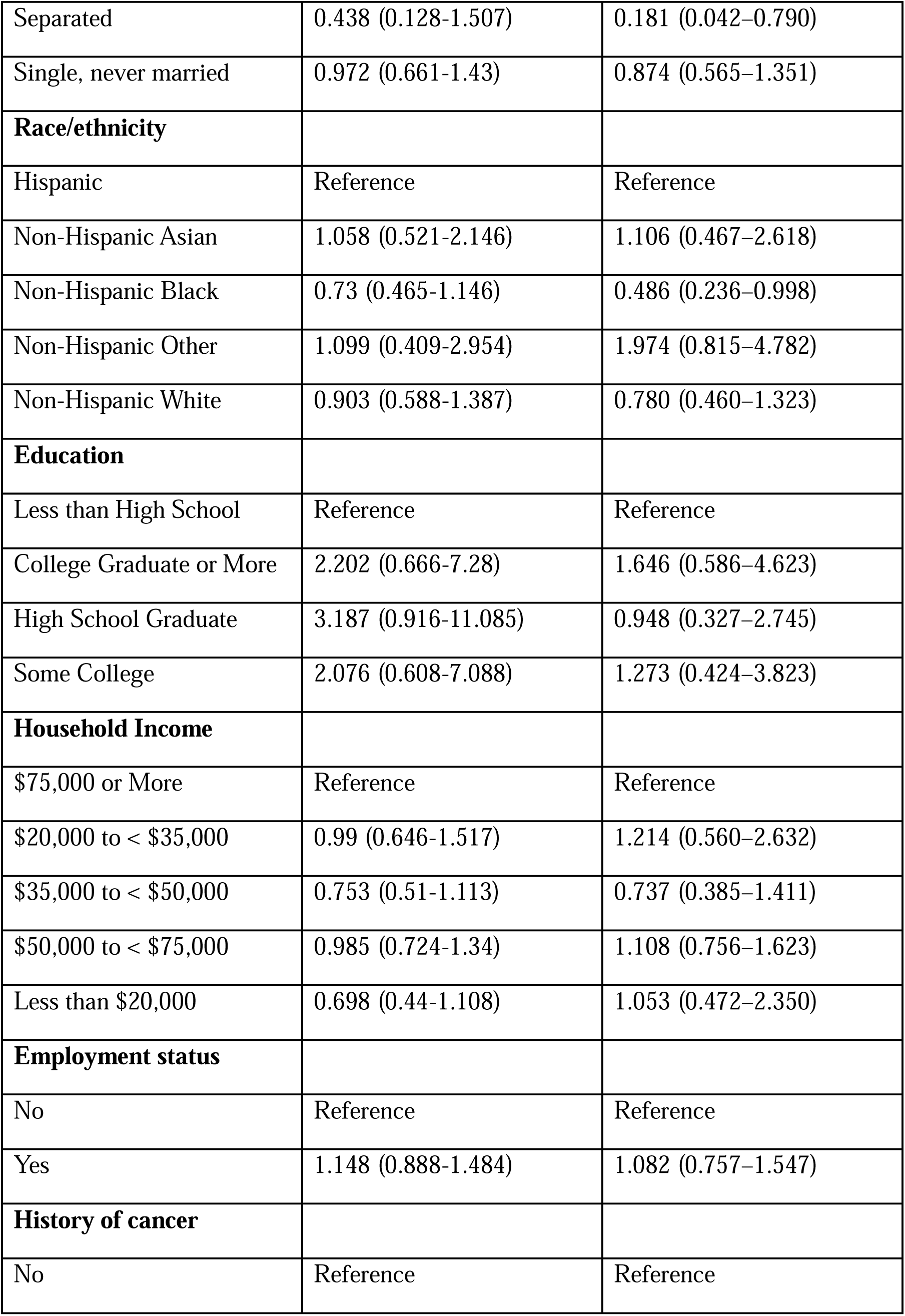
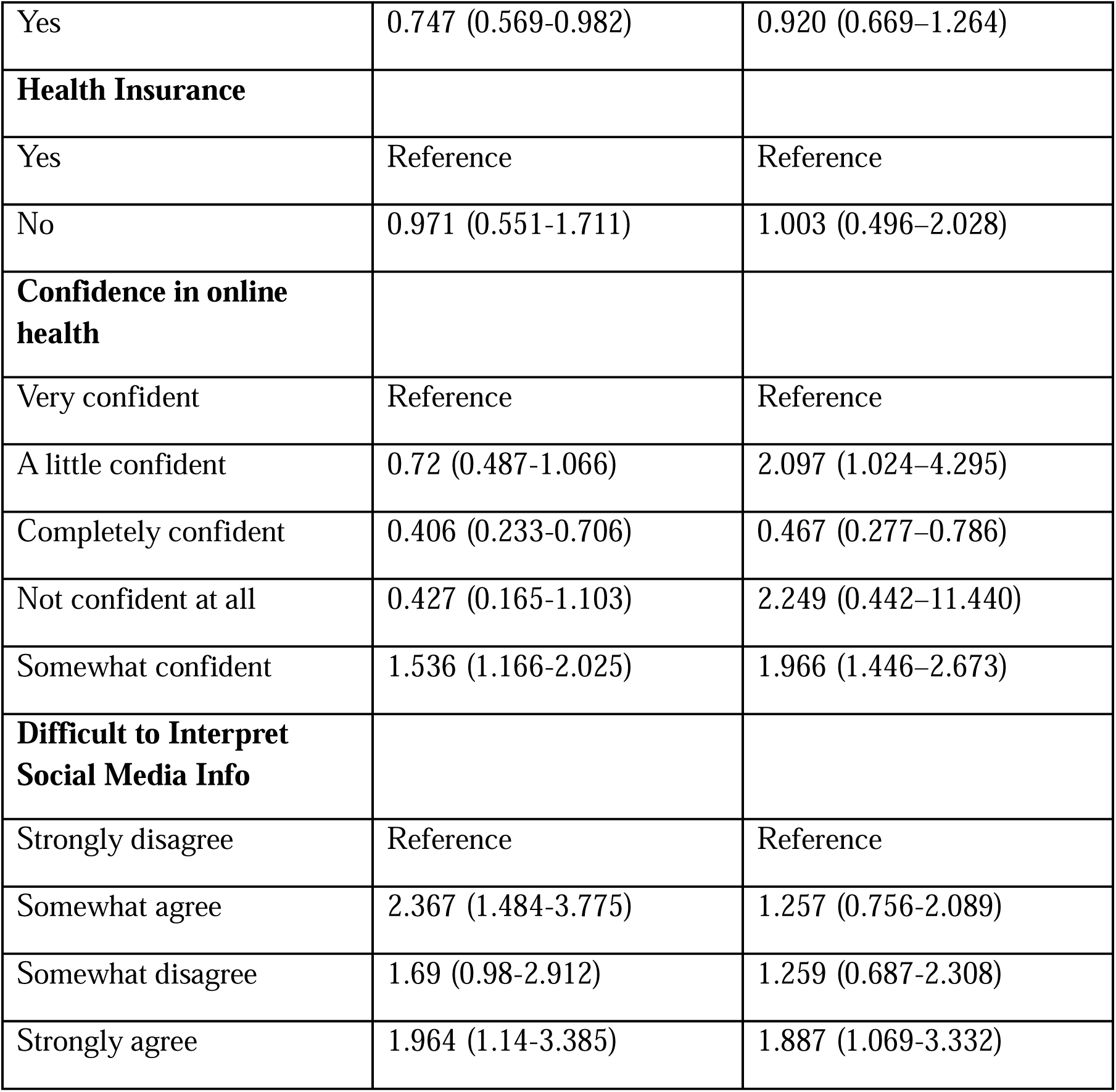
Univariate and Adjusted Multivariable Regression Analysis of the Association between Concern about Cancer Info Quality, and Participant Characteristics.

Individuals having difficulties in understanding medical statistics had higher odds (AOR=1.889; 95% CI: 1.17-3.048) of interpreting the cancer related information compared to the complementary group.

The way people understand cancer-related information shows an interesting pattern. Respondents who felt ‘completely confident’ in finding health-related information on the internet had significantly lower odds (AOR=0.564; 95% CL: 0.324-0.983) of having difficulty in understand cancer related information compared to the respondents who were ‘very confident’.

Conversely, people who demonstrated moderate self-assurance (‘somewhat confident’) about their knowledge actually performed worse than those who were very confident in grasping the knowledge on cancer related information. The odds of difficulty were approximately twice as high (AOR=2.074; 95% CL:1.404-3.063) compared to respondents who were ‘very confident’ and more than three times higher (AOR=3.059; 95% CL: 1.493-6.266) for those who described it as ‘a little confident’. The results show that even a small amount of self-doubt can make it difficult to understand complex cancer information which demonstrates why confidence matters when dealing with these complicated subjects.

Furthermore, respondents who strongly agreed on facing difficulty telling if health related information found on social media is true or false had higher odds (AOR:2.924; 95% CL: 1.664-5.139) of having difficulty in understanding the cancer related information than those who disagreed strongly.

Among other predictors, widowed individuals demonstrated significantly higher odds (AOR=3.026; 95% CL:1.622-5.647) of difficulty in comprehending cancer-related information compared to their married counterparts, while those with household incomes between $50,000 and $75,000 showed higher odds (AOR: 1.6; 95% CI: 1.023-2.503) of difficulty in understanding the cancer information relative to individuals earning $75,000 or more.”

For Cancerconcernedquality: Respondents whose perception of understanding medical statistics is hard, is associated with significantly higher odds (AOR=1.691; 95% CI:1.113–2.570) of concern about cancer quality. On the other hand, elderly individuals aged 75 years and above and those who report being in separated marital status have significantly lower odds (AOR=0.500; 95% CL: 0.294–0.851), (AOR=0.181; 95% CL: 0.042–0.790) than their corresponding reference groups. In terms of race/ethnicity, non-Hispanic black respondents have significantly lower odds (AOR= 0.486; 95% CL: 0.236–0.998) of concern than Hispanic individuals, while other racial/ethnic groups did not differ significantly from the reference category.

Educational attainment was not significantly associated with concern about cancer information quality in the adjusted model, though higher proportions of college-educated respondents expressed concern in the univariate analysis. Similarly, household was not significantly associated with concern, although individuals with annual incomes between $50,000 and $75,000 exhibited slightly elevated odds compared to those earning $75,000 or more. Work status also did not show any statistically meaningful differences between full-time workers and non-full-time workers.

Finally, regarding information found digitally, ‘completely confident’ respondents have significantly lower odds (AOR=0.467; 95% CL: 0.277–0.786) of concern than those ‘Somewhat confident’ with odds (AOR=1.966; 95% CL: 1.446–2.673). Additionally, those who were a little confident also had elevated odds (AOR=2.097; 95% CI: 1.024–4.295) relative to the very confident group. Respondents who strongly agree on questioning the credibility of health-related information found on social media, are associated with higher odds (AOR=1.887; 95% CL: 1.069-3.332) of concern on cancer related information.

## Discussion

This study examined the relationship that exists between the perceived challenge in reading medical statistics and the way people perceive information about cancer. Two notable trends were to be observed. To begin with, respondents who struggled with medical statistics were highly likely to report increased concern regarding the quality of information on cancer and struggle to understand it. Second, the use of social media as a source of health information was closely linked to the negative perceptions in both areas. These findings collectively highlight the dual impact of statistical literacy and information environment in shaping how the public interprets and evaluates cancer communication.

Our results expand and extend prior research on health literacy and communication disparities. Earlier studies have shown that limited health literacy undermines the ability to process complex health information and corrode confidence in medical decision-making (Sørensen et al., 2012; Halverson et al., 2015). Similarly, Holden et al. (2021) demonstrated that lower numeracy and literacy predict not only reduced comprehension but also underprivileged health-related quality of life. In line with this evidence, our findings confirm that comprehension and perceptions of quality are interrelated challenges rather than distinct outcomes.

The strong association between social media use and negative perceptions reflects a growing body of research on the impact of digital information environments. While online platforms expand access to health information, they also increase exposure to misinformation, contradictory claims, and unverified content. Recent studies have emphasized that individuals who rely heavily on social media may experience heightened uncertainty and distrust, especially when information sources appear inconsistent or lack credibility. Our study adds to this literature by showing that social media use is not only associated with concerns about information quality but also with the perceived difficulty of understanding cancer content itself, suggesting a deeper interaction between information environment and comprehension.

Health literacy, defined as “the degree to which individuals can obtain, process, understand, and communicate about health-related information needed to make informed health decisions” (Berkman et al., 2010) is a complex, multifaceted phenomenon.

In the health litreacy framework, communication is structured around four stages: (1) Access - easy access to information about screening; (2) Understand- use of plain language that is easy to comprehend; (3) Appraise- frame information in a way so that individuals can judge its relevance; (4) Apply- clarifying the next steps and minimize barriers to taking it (President’s Cancer Panel, 2022).

There are multiple ways to raise awareness of a disease like Cancer to reduce health risks, and to better support people who have been suffering from it. The initiative The Cancer Health Awareness through screening and Education (CHANGE) (Vernon, et.al, 2024) was conceived to improve cancer care and reduce racial disparities and inequities in a sustainable and collaborative community based manner by: (1) increasing evidence based cancer awareness though health literacy and education, with an emphasis on prevention of modifiable risk factors, screening, and early detection; and (2) providing access and navigation to high quality cancer screening and early detection services. Pre- and post-intervention knowledge, attitudes and beliefs were measured, as well as changes in cancer health behaviors and rates of screening.

Cancer risk communication can be difficult to understand for many audiences because the concepts are probabilistic, evolving and use jargons. Individuals who do not work in health care or quantitative fields, may have limited health literacy and numeracy relative to those demands. This instance reflects a message–audience mismatch rather than a fault from the recipient’s end (Fischhoff, 1999).

According to the Vintage 2024 Population Estimates released by the U.S. Census Bureau, the share of U.S. residents aged 65 and above, shows an upward trend from 12.4% in 2004 to 18.0% in 2024, while the children cohort declined from 25.0% to 21.5%. In our bivariate analysis, we found the elderly-particularly aged 75 years or above, those living alone, may show fewer quality concerns and less likely to share opinions whether cancer information is “hard to understand”, which indicates at: a) lower exposure to high noise digital channels leading to absence of doubts or questions, b)inability to access health information on internet due to lack of being tech savvy, c) lack of vision, working memory, and numeracy due to aging can increase the cognitive effort required to comprehend the information found online and medical statistics. Health care communicators should incorporate these considerations while designing messages and interventions that encourage active engagement among older adults in cancer communication (Sparks & Nussbaum, 2008). Social media can be an effective medium to raise awareness of cancer symptoms, encourage help seeking, and attempting to influence social norms around help seeking via national public health campaigns to improve early diagnosis of cancer. That being said, effects are often short-lived, predominantly one-sided and unevenly distributed with limited benefit for those most are in need (Plackett et al., 2020).

Additionally, Widowed respondents exhibited higher odds of reporting difficulty understanding cancer information and participants with a household income of $50–$75k were more likely to find cancer related information hard to understand, compared to those with incomes >$75k. Given the older adults living alone - including widowed individuals bear an elevated cancer risk, awareness and health numeracy education efforts, led by health-care professionals and national/state public health agencies are likely to improve comprehension and screening uptake.

There are a few limitations that should be considered. To begin, the cross-sectional design does not allow making causal conclusions, and it is possible that the people who maintain distrust towards cancer information are simply less willing to pursue or attain statistical literacy. Second, self-reports are also subject to recall and reporting bias, because people have the possibility of over- or underestimating their comprehension capacity. Third, although our models have accounted a variety of demographic and behavioral factors, unmeasured variables like previous cancer experience, cultural beliefs or even general numeracy may affect perceptions. Lastly, the use of survey data restricts the possibility of immersing into the intricate ways in which people communicate with statistical and digital material in the real world. Further studies are needed to investigate longitudinal designs in order to measure the changes in the status of statistical literacy and the perception of cancer information over time and the impact of specific interventions. The experimental research to test the usefulness of simplified statistical communication strategies would aid to find some methods of enhancing their understanding and credibility. Moreover, qualitative research, including focus groups or interviews, can be used to gain an understanding of how people cope with conflicting messages about cancer on various platforms, especially on social media. Since the role of digital health communication is currently increasingly important, it would also be worth exploring platform-specific variables, including algorithms curation, statistics visualizations, and message framing. Lastly, research evaluating the effectiveness of interventions to improve statistical literacy in diverse groups of people could help explain how specific measures can decrease cancer communication disparities.

## Conclusion

In conclusion, this study highlights the pivotal nature of statistical literacy as a key factor that helps to develop the perspective of the population regarding the information about cancer. The problem in interpreting medical statistics was highly associated with two issues: quality concerns and the feeling of incomprehensibility. Collectively, these results point to the fact that understanding and trust are not independent phenomena. As a public health concern, it is more than just increasing access to information that will resolve these problems. Individuals also need skills to analyze medical statistics as well as be able to critically review digital health information. Enhancing the enhancement of statistical literacy, accompanied by enhancing the comprehensibility and believability of cancer-related communication on the Internet, will be essential towards making informed decisions, reducing disparities, and increasing trust in cancer information.

## Acknowledgements

The authors thank the National Cancer Institute for providing public access to the 2022 Health Information National Trends Survey (HINTS) dataset used in this secondary analysis.

## Ethics Statement

The study used publicly available, de-identified data from the Health Information National Trends Survey (HINTS) and does not require additional ethical approval from author’s Institutional Review Board

## Informed Consent

Not applicable

## Data Availability Statement

The data used in this study is publicly available from the Health Information National Trends Survey (HINTS) 2023 survey. The 2022 HINTS data files and accompanying documentation can be accessed through the HINTS website (https://hints.cancer.gov/data/download-data.aspx#H6).

## Reference

Finney Rutten, L. J., Blake, K. D., Greenberg-Worisek, A. J., Allen, S. V., Moser, R. P., & Hesse, B. W. (2019). Online Health Information Seeking Among US Adults: Measuring Progress Toward a Healthy People 2020 Objective. Public health reports (Washington, D.C.: 1974), 134(6), 617–625. 10.1177/0033354919874074

Liu, P. L., & Jiang, S. (2021). Patient-Centered Communication Mediates the Relationship between Health Information Acquisition and Patient Trust in Physicians: A Five-Year Comparison in China. Health Communication, 36(2), 207–216. 10.1080/10410236.2019.1673948

Bennett, I. M., Chen, J., Soroui, J. S., & White, S. (2009). The contribution of health literacy to disparities in self-rated health status and preventive health behaviors in older adults. Annals of Family Medicine, 7(3), 204–211. 10.1370/afm.940

Viswanath, K., & Kreuter, M. W. (2007). Health disparities, communication inequalities, and eHealth. American Journal of Preventive Medicine, 32(5 Suppl), S131–S133. 10.1016/j.amepre.2007.02.012

Tomar, A., Harvey, I., Meng, X., & Feng, S. (2024). Using structural equation modelling to explore the relationship between patient-centered communication, human papilloma (HPV)-related knowledge, and perceived effectiveness of the HPV vaccine. Asian Pacific Journal of Cancer Prevention, 25(8), 2761–2772. 10.31557/APJCP.2024.25.8.2761

Zaidi, M., Sarkar, S., Arakelyan, S., & Poghosyan, H. (2024). Relationship between fatalistic cancer beliefs and risky health behaviors. Western Journal of Nursing Research, 46(10), 757–765. 10.1177/01939459241273388

Elkefi, S., & Matthews, A. K. (2024). Exploring health information-seeking behavior and information source preferences among a diverse sample of cancer survivors: Implications for patient education. Journal of Cancer Education, 39(6), 650–662. 10.1007/s13187-024-02448-3

Ezeani, A., Boggan, B., Hopper, L. N., Herren, O. M., & Agurs-Collins, T. (2023). Associations between cancer risk perceptions, self-efficacy, and health behaviors by BMI category and race and ethnicity. International Journal of Behavioral Medicine. Advance online publication. 10.1007/s12529-023-10225-7

Allen, C. G., Green, R. F., Dowling, N. F., Fairley, T. L., & Khoury, M. J. (2023). Understanding the process of family cancer history collection and health information seeking. Health Education & Behavior, 50(5), 572–585. 10.1177/10901981231152430

Van Heel, K. L., Nelson, A., Handysides, D., & Shah, H. (2023). The factors associated with confidence in using the internet to access health information: Cross-sectional data analysis. JMIR Formative Research, 7, e39891. 10.2196/39891

MacKay, E. N., & Sellers, A. H. (1970). A prospective trial of the TNM classification of breast cancer by the regional cancer treatment centres in Ontario, 1960–1964. International Journal of Cancer, 6(3), 517–528. 10.1002/ijc.2910060323

Kessler, D. A., Escamilla, L. A., Pan, S., & Di Valentino, E. (2025). One-parameter dynamical dark energy: Hints for oscillations. arXiv Preprint, arXiv:2504.00776. https://arxiv.org/abs/2504.00776

Garcia-Retamero, R., & Cokely, E. T. (2017). Designing Visual Aids That Promote Risk Literacy: A Systematic Review of Health Research and Evidence-Based Design Heuristics. Human factors, 59(4), 582–627. 10.1177/0018720817690634

Lipkus, I. M., & Peters, E. (2009). Understanding the role of numeracy in health: proposed theoretical framework and practical insights. Health education & behavior: the official publication of the Society for Public Health Education, 36(6), 1065–1081. 10.1177/1090198109341533

Galesic, M., & Garcia-Retamero, R. (2011). Graph literacy: a cross-cultural comparison. Medical decision making: an international journal of the Society for Medical Decision Making, 31(3), 444–457. 10.1177/0272989X10373805

Holden, C. E., Wheelwright, S., Harle, A., & Wagland, R. (2021). The role of health literacy in cancer care: A mixed studies systematic review. PloS one, 16(11), e0259815. 10.1371/journal.pone.0259815

Berkman, N. D., Davis, T. C., & McCormack, L. (2010). Health Literacy: What Is It? Journal of Health Communication, 15(sup2), 9–19. 10.1080/10810730.2010.499985

President’s Cancer Panel. (2022). Closing gaps in cancer screening: Connecting people, communities, and systems to improve equity and access. U.S. Department of Health and Human Services, National Cancer Institute. https://prescancerpanel.cancer.gov/reports-meetings/cancer-screening-report-2022

Vernon, M., Coughlin, S. S., Tingen, M., Jones, S., & Heboyan, V. (2024b). Cancer health awareness through screening and education: A community approach to healthy equity. Cancer Medicine, 13(13), e7357. 10.1002/cam4.7357

Fischhoff, B. (1999). Why (Cancer) risk Communication can be hard. JNCI Monographs, 1999(25), 7–13. 10.1093/oxfordjournals.jncimonographs.a024213

Sparks, L., & Nussbaum, J. F. (2008). Health literacy and cancer communication with older adults. Patient education and counseling, 71(3), 345–350. 10.1016/j.pec.2008.02.007

Plackett, R., Kaushal, A., Kassianos, A. P., Cross, A., Lewins, D., Sheringham, J., Waller, J., & Von Wagner, C. (2020). Use of social media to promote cancer screening and early diagnosis: Scoping review. Journal of Medical Internet Research, 22(11), e21582. 10.2196/21582

Abdelhadi O. (2023). The impact of psychological distress on quality of care and access to mental health services in cancer survivors. Frontiers in health services, 3, 1111677. 10.3389/frhs.2023.1111677

Sørensen, K., Van den Broucke, S., Fullam, J., Doyle, G., Pelikan, J., Slonska, Z., Brand, H., & (HLS-EU) Consortium Health Literacy Project European (2012). Health literacy and public health: a systematic review and integration of definitions and models. BMC public health, 12, 80. 10.1186/1471-2458-12-80

Halverson, J. L., Martinez-Donate, A. P., Palta, M., Leal, T., Lubner, S., Walsh, M. C., Schaaf Strickland, J., Smith, P. D., & Trentham-Dietz, A. (2015). Health Literacy and Health-Related Quality of Life Among a Population-Based Sample of Cancer Patients. Journal of health communication, 20(11), 1320–1329. 10.1080/10810730.2015.1018638

